# Association of 25-OH Vitamin D Levels With In-Hospital Mortality: A cross-sectional, observational study of the MIMICIV database

**DOI:** 10.1101/2024.06.05.24308492

**Authors:** Yaotang Wang, Weijing Lv, Haixia Yu

## Abstract

The relationship between 25-OH Vitamin D3 and hospital death continues to be a topic of controversy. At present, the available evidence regarding the relationship between 25-OH Vitamin D and hospital death is inadequate. Therefore, we conducted a cross-sectional study involving 197 sepsis-3 participants from MIMIC-IV. Basic information, laboratory indicators, underlying Disease, therapeutic medications and tools, disease score were obtained from all participants. Serum 25-OH Vitamin D was collected on Intensive Care Unit (ICU) admission, and inpatient mortality were analyzed. The logistic regression, fitting curve and interaction effects were utilized to substantiate the correlation of serum 25-OH Vitamin D with inpatient mortality. Furthermore, interaction and subgroup analyses were evaluated for the consistency of these correlations. A total of 197 patients were included in the analysis, with an average age of 59.8 years and 43.1% male. The mean 25-OH vitamin D level in the cohort was 21 ng/mL. We observed no significant association between higher 25-OH vitamin D levels and increased inpatient mortality rates. The in-hospital mortality did not differ among patients vitamin D deficiency, insufficiency and sufficiency group (15.7% vs 17.4% vs 24.5%, p=0.417). A multivariable adjusted logistic regression model showed that vitamin D levels could not predict in-hospital mortality (OR 1.02 95% CI: 0.98∼1.05, p=0.342, Model 3). In conclusion, 25-OH vitamin D detected upon ICU admission was not associated with in-hospital mortality in patients with sepsis.

## Introduction

Sepsis is a serious condition caused by the body’s abnormal response to an infection, resulting in life-threatening organ dysfunction(1). It is a leading cause of death in intensive care unit (ICU) patients, and its mortality rates have increased to over 30% due to factors such as global aging, invasive surgeries, immunosuppressant usage, and the rise of antibiotic-resistant bacteria(2). To address this, it is crucial to study the factors that impact sepsis mortality in order to reduce the mortality rate.

Vitamin D, a fat-soluble hormone, plays a crucial role in regulating calcium and phosphorus levels, maintaining bone and cardiovascular health(3), and boosting immunity(4).

Vitamin D is an essential nutrient for humans that was discovered in 1921. In addition to regulating calcium and phosphate metabolism, it is important for immunity, endothelial function, and antimicrobial activity(5). Previous related studies have evaluated the relationship between vitamin D deficiency and mortality risk in patients with sepsis. A two-center observational study said that 25-OH vitamin D deficiency prior to hospital admission is a significant predictor of sepsis in the critically ill, which has an increased risk of mortality following critical care initiation(6). While, a prospective multicentre observational study illustrated that 25-OH vitamin D deficiency detected upon ICU admission was not associated with 90-day mortality in patients with severe sepsis(7).

It is unclear whether lower serum 25 (OH) D predicts poor outcome in sepsis. Therefore, we aimed to conduct an association analysis of the MIMIC data to comprehensively assess the relationship between serum 25 (OH) D and the risk of in-hospital mortality in adult patients with sepsis.

## Materials and Methods

### Data Source

We performed a retrospective cohort study using the Medical Information Mart for Intensive Care (MIMIC-IV) (version 2.2)(8,9), which is a large publicly accessed database. The use of this database has been approved by the Massachusetts Institute of Technology and the Institutional Review Board of Beth Israel Deaconess Medical Center (BIDMC, Boston, MA, USA). One of the authors, Yaotang Wang, accomplished the National Institutes of Health’s web-based course “Data or Specimens Only Research”(Record ID: 59979404) and “Conflicts of Interest”(Record ID: 59979406) and was approved to access the database to extract data.

To protect patient privacy, all data were de-identified. Thus, informed consent was waived by the ethical committee of the Beth Israel Deaconess Medical Center. This study is described in conformity to the strengthening the Reporting of observational studies in Epidemiology (STROBE) statement and was managed to conform to the tenets of the Declaration of Helsinki.

### Study Population and Variable Extraction

The total number of patients in the MIMIC-IV included 299,712 individuals from 2008 to 2019, of which 73,181 were admitted to the ICU. Among them, 25647 patients with sepsis-3 were selected based on definition(10). Patients >18 years were initially enrolled in this study, and only data for the first hospitalization and ICU stay were collected for patients. Meanwhile, patients without serum 25-OH-VitaminD after ICU admission who stayed in the ICU were excluded. Thus, only 197 patients were included in this study after further excluding covariates with missing values(Figure 1). The first test serum 25-OH-VitaminD after ICU admission was extracted as the interest variable and the major exposure factor in this study. All variables in this study were extracted from the MIMIC-IV database using Structured Query Language (SQL) with PostgreSQL. Demographic variables, including age, sex, ethnicity and marital status, were obtained. Clinical severity on admission was examined using the APACHE II score and SOFA score. Vital signs in this study also included, such as temperature, heart rate, respiratory rate (RR), and percutaneous oxygen saturation (SpO2). Comorbidities, including myocardial infarction, congestive heart failure, chronic pulmonary disease, renal failure, diabetes without complication, diabetes with complication and renal disease and malignant cancer, were also included for analysis based on the recorded ICD-9 and ICD-10 codes from the database. Laboratory variables, including hematocrit, platelet count, white blood cell (WBC) count, anion gap, bicarbonate, sodium, potassium, glucose, chloride, creatinine and blood urea nitrogen (BUN), were obtained within the first test after ICU admission. The details of the missing value are shown in Table 1.

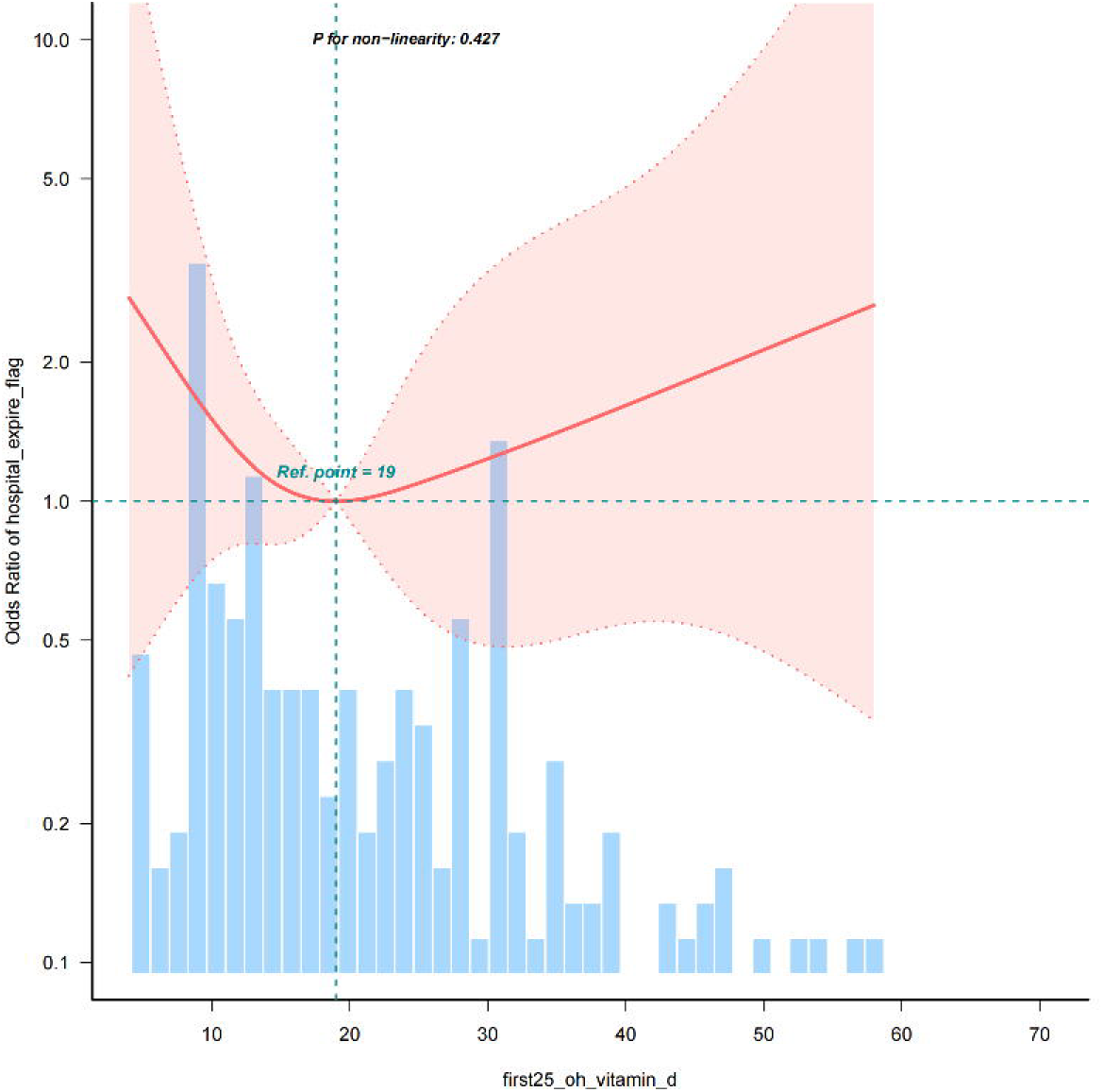

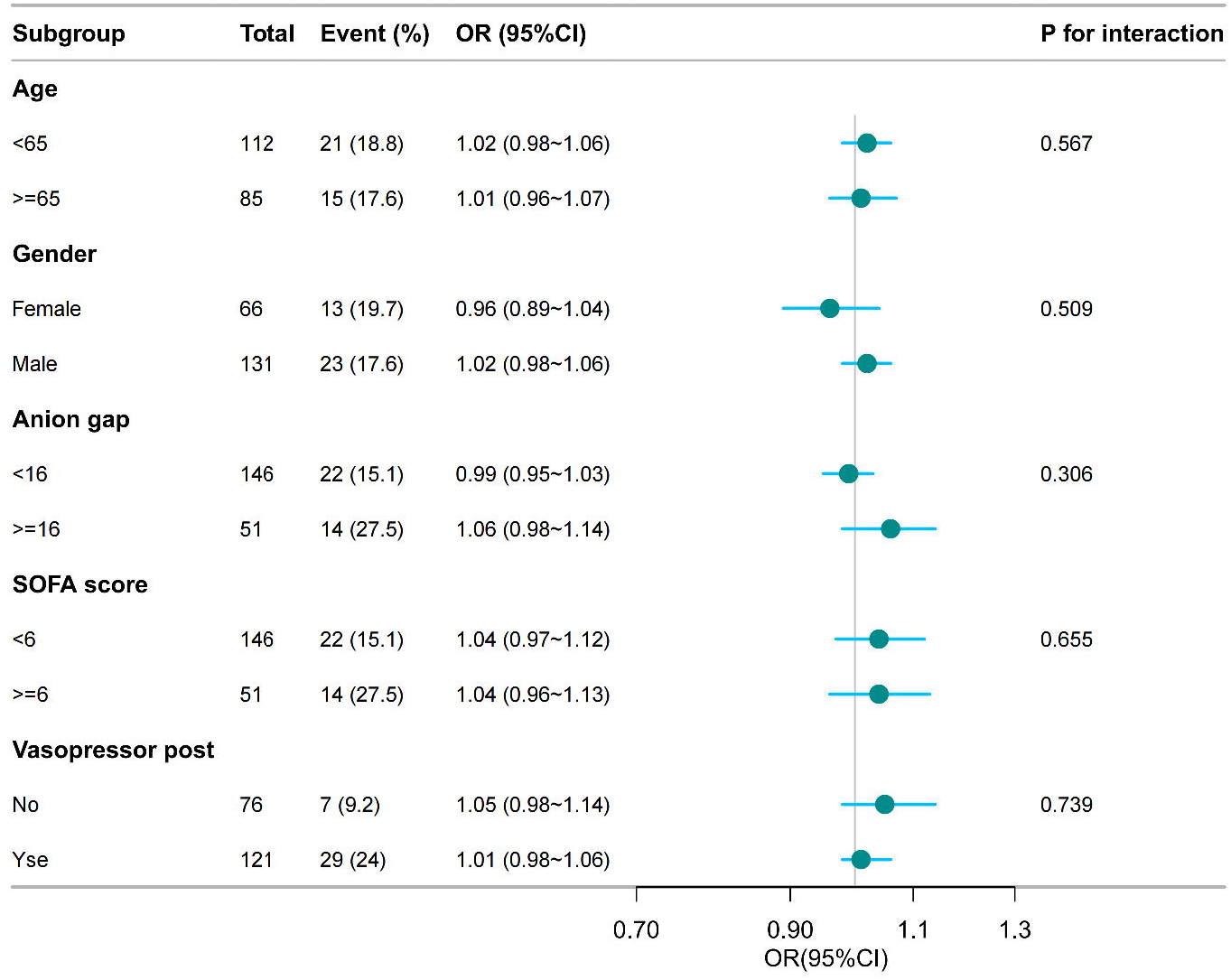

### Outcomes

The primary endpoint was hospital inpatient death, which was defined by patient survival status at the time of hospital.

### Statistical analysis

Continuous variables were described as means±standard deviation (SD) or as median interquartile ranges (IQR), and categorical variables were described as percentages. We used Fisher’s exact and Chi-square tests or the Kruskal-Wallis test to examine the statistical differences among three groups—the deficient serum 25-OH-VitaminD (AG <19.9ng/ml), insufficient serum 25-OH-VitaminD(20ng/ml≤25-OH-VitaminD<30ng/ml) and sufficient 25-OH-VitaminD(> 30ng/ml)groups(11). We used multivariate logistic regression models to evaluate the relationship of serum 25-OH-VitaminD and inpatient mortality. Restricted cubic spline analysis was used to describe the non-linear association between serum 25-OH-VitaminD and inpatient mortality.

Baseline variables that were considered clinically relevant or had a change in effect estimate of >10% were chosen as confounders. In Model I, the covariates were adjusted for age,gender,temperature,SpO2,marital status,potassium. As for adjusted model, the covariates were additionally included with mycardial infraction,diabetes with complications and vasopressors post.

Furthermore,interactions and stratified analyses were conducted using age,gender,temperature,SpO2,marital status,potassium,mycardial infraction,diabetes with complications and vasopressors post, as previously described. For the missing data, we directly excluded them. All results were expressed as odds ratios (OR) with a 95% CI, and a p-value of <0.05 was considered significant. All analyses were performed using the statistical software packages R 3.3.2 (http://www.R-project.org, The R Foundation) and Free Statistics software version 1.9.2 (Beijing, China)(12).

## Results

### Baseline Characteristics of Subjects

In total, 216 of the 25647 sepsis-3 patients who received 25-OH-VitaminD measurement were included in this study (Figure 1). Excluding those with missing some covariates, a total of 197 people were included in the final study. Among them, there were 85 males and 112 females, and the mean age was 59.8 ± 16.9. The distribution of the baseline population characteristics according to serum 25-OH-VitaminD levels in tertiles is described in Table 2. The patients with sufficient serum 25-OH-VitaminD levels (≥30ng/ml) were found to have a higher age and bicarbonate, a lower anion gap and creatine level.

### Association between 25-OH Vitamin D Levels and inpatient mortality

Table 2 presents the results of the multivariable logistic regression analysis examining the association between 25-OH-VitaminD and inpatient death. A high 25-OH-VitaminD wasn’t associated with an increased risk of inpatient death (Per 1 unit: OR=1.02, 95% CI=0.98∼1.05, P=0.342), after adjusting for potential confounders (Table 2, all adjusted model). When 25-OH-VitaminD was divided into three tertiles, multivariable-adjusted regression odds ratio (95% CIs) of 25-OH-VitaminD associated with based on 25-OH-VitaminD of group1 and group2 was 0.76 (95% CI=0.29∼1.99, p=0.574), and 0.75 (95% CI=0.25∼2.23, p=0.601) when comparing the 25-OH-VitaminD sufficient group, respectively (Table 2, all-adjusted model).

Evidence from the estimated dose-response curve indicated a linear relationship between 25-OH Vitamin D and risk of inpatient death (Figure 3, P for non-linearity =0.176, restricted cubic spline analyses adjusted for multiple variables, and the highest and lowest 0.5% was trimmed for each 25-OH Vitamin D measure.)

### Subgroup Analysis

Subgroup analyses were performed to evaluate the association between serum 25-OH Vitamin D levels and inpatient mortality (Figure 4) among subgrs. Based on the variables tested, an association between higher serum 25-OH Vitamin D level and inpatient mortality was observed in different subgroups (age, gender,anion gap, sofa score and vasopressor post) (p >0.05 for all).

## Discussion

Vitamin D is involved in the inflammatory response and in the regulation of the body’s immunity.There are many conflicting research opinions on the correlation between vitamin D and sepsis mortality, with some studies suggesting that lower vitamin D levels are associated with increased mortality in sepsis patients. However, there have been other studies that have found no significant effect of vitamin D supplementation on survival in sepsis patients. Therefore, while there are some studies that support an association between vitamin D and sepsis mortality, this is an area where further research is needed to draw definitive conclusions.

The mechanism by which vitamin D deficiency is associated with sepsis death may be due to an increased susceptibility to sepsis caused by the dysfunction of immune cells as well as barrier cells(13). On the other hand, the reduction of 1,25 (OH) 2D3 at the tissue level may reduce the body’s ability to regulate autoimmunity and affect the expression of antimicrobial peptides(14), affect the body’s innate immune regulation, and further increase the sepsis susceptibility(15).

Two meta-analyses from 2014 and 2015 suggested that vitamin D deficiency increased susceptibility for sepsis(16,17). A prospective study of 88 patients showed that low serum vitamin D level on admission (<8.1 ng/mL) was associated with increased 28-day mortality in sepsis, and multivariate COX regression analysis showed that vitamin D concentrations <8.1 ng/mL (p=0.01) and septic shock (p=0.02) were associated with increased mortality in sepsis. The hazard ratio for 28-day mortality with a vitamin D level <8.1 ng/mL was 1.95 (95% CI 1.15 – 3.29, p=0.01, and the mean survival of patients with vitamin D levels <8.1 ng/mL was 9 days (median 6 days), while those with vitamin D levels of 8.1 ng / mL had a mean survival of 14 days (median 10 days)(18). A study of the role of 1,25-dihydroxyvitamin d3 in experimental sepsis showed no regulation of vitamin D in lipopolysaccharide-induced sepsis(19).A study of 116 cases on the relationship between vitamin D and COVID-19 inflammation storm showed that high-dose vitamin D is considered a promising treatment to inhibit cytokine storms in COVID-19 patients, and is associated with better clinical improvement and less adverse outcomes(20).An observational study showed that 107 critically ill adult patients with sepsis reported vitamin D deficiency in 93.5% of patients, and 53.3% of them showed extremely low 25 (OH) D flow concentration (7 ng/mL), specifically, extremely low 25 (OH) D flow concentration and higher mean SAPS 2 score were independent predictors of sepsis-related mortality(21).

Compared with the previous study, our study reached a different conclusion, our results show: in multivariate regression analysis, in vitamin D sufficient group, insufficient group, and lack group, unadjusted model, model 1 and full model respectively, shows that VD and in-hospital mortality no curve relationship and multivariate analysis without statistical significance, VD is neither dangerous nor protective factors, and model OR changes between each group, the model is robust.

Although most studies showed a significant inverse association between vitamin D level and mortality,there are also previous studies consistent with our findings. A single-center prospective study showed that vitamin D was not associated with in-hospital mortality(22). An additional prospective study of 461 people analyzing the association of 25 (OH) D and 1,25 (OH) D with sepsis mortality also showed that vitamin D did not predict death in patients with sepsis(23).

In addition, the epidemiological results of our study showed that the total number of 25-OH-VitaminD (<19.9) and 25 (2025-OH-VitaminD <30) (51%, 23%) and 49 (30 25-OH-VitaminD) in the adequacy group 49 (24%). The mortality rate in the VD deficiency group was 15.7% versus 24.5%, respectively. The difference between 25-OH-VitaminD deficiency was insignificant (15.7VS17.4% VS 24.5%, p=0.417). Accordingly, a prospective observational study conducted by Konlawij Trongtrakul et al had expressed that among septic patients the total number of 25-OH-VitaminD deficient (<20 ng/mL) and non-deficient (≥20 ng/mL) was 110, or 83,27 (75%, 25%, respectively). The mortality rate in the VD deficient and non-deficient groups were 16% versus 4%, respectively. The total number of patients (<12 ng/mL) (12 ng/mL) was 53,57, respectively, mortality in the VD deficiency group was 23% versus 4% in the non-severe deficiency group, respectively, 30-day in-hospital mortality was not significantly higher in the vitamin D deficiency group (16% vs 4%, p=0.18), but the difference between the severe deficiency group and the non-severe group was significant (23% vs 4%, p=0.02)(25). Compared with the similar studies, the vitamin d deficiency groups in the two studies showed similar mortality (15.7% VS 16%), but not in deficiency (20 ng/ml) (10% VS 4%). In our study, the population in the vitamin D non-deficiency group, mortality was higher than that in the other study populations and no significant relation among tertiles were found.

The reasons for the above results may be as follows: First, the sepsis patients admitted to ICU are in critical condition, and after a period of poor treatment effect, they eventually develop critically ill. As a result, most of the patients are in bed or semi-bed state, with insufficient light, poor diet and decreased absorption capacity.Second, most of the patients in ICU were septic shock. On the first day of admission, after a large amount of fluid resuscitation treatment, dilutional factors led to the relative reduction of vitamin D. The underlying causes for these findings could be: Initially, patients with sepsis in the ICU are in a critical state, and following a phase of ineffective treatment, they ultimately become critically ill. Consequently, a majority of the patients remain in a bed or semi-bed condition, suffering from inadequate lighting, a substandard diet, and reduced absorption ability. A majority of ICU patients experienced septic shock. Upon initial hospital admission, following extensive fluid resuscitation, factors causing dilution resulted in a noticeable decrease in vitamin D levels. Additionally, the observational design of the study meant that its findings could only indicate a correlation, not a direct cause-and-effect link. Thirdly, there could be complicating elements that the multivariate regression analysis overlooked.

Overall, our study produced negative results, probably because the vitamin D and sepsis and related covariates in MIMIC-IV database, only 197 people, sample size factors, of course, there are bias in retrospective cohort study, further for the large sample population and multicenter study, look forward to further clarify the correlation between vitamin D and sepsis patients mortality, hope early detection of sepsis patients and reduce the incidence and mortality of severe patients.

## Conclusions

In conclusion, vitamin D deficiency at ICU admission was not associated with in-hospital mortality in sepsis patients.

## Data Availability

The MIMIC data analyzed in the current study are publicly available.

## Data Availability

The MIMIC data analyzed in the current study are publicly available.

## Acknowledgments

We would like to thank the Massachusetts Institute of Technology and the Beth Israel Deaconess Medical Center for the MIMIC-IV database.We thank Dr. Jie Liu of the Department of Vascular and Internal Medicine, People’s Liberation Army General Hospital, China, for his assistance. Dr. Jie Liu, Department ofVascular and Internal Medicine, People’ s Liberation Army General Hospital, China, for his contributions to statistical support, study design consultation, and comments on the manuscript.

## Supplementary Materials

Figures and tables are separately unloaded on dashboard.

